# Clinicopathological Heterogeneity of Lewy Body Diseases: The Profound Influence of Comorbid Alzheimer’s Disease

**DOI:** 10.1101/2024.08.30.24312864

**Authors:** Thomas G. Beach, Geidy E. Serrano, Nan Zhang, Erika D. Driver-Dunckley, Lucia I. Sue, Holly A. Shill, Shyamal H. Mehta, Christine Belden, Cecilia Tremblay, Parichita Choudhury, Alireza Atri, Charles H. Adler

## Abstract

In recent years, proposals have been advanced to redefine or reclassify Lewy body disorders by merging the long-established entities of Parkinson’s disease (PD), Parkinson’s disease dementia (PDD) and dementia with Lewy bodies (DLB). These proposals reject the International DLB Consortium classification system that has evolved over three decades of consensus collaborations between neurologists, neuropsychologists and neuropathologists. While the Consortium’s “one year rule” for separating PD and DLB has been criticized as arbitrary, it has been a pragmatic and effective tool for splitting the continuum between the two entities. In addition to the decades of literature supporting the non-homogeneity of PD and DLB, it has become increasingly apparent that Lewy body disorders may fundamentally differ in their etiology. Most PD subjects, as well as most clinically-presenting DLB subjects, might best be classified as having a “primary synucleinopathy” while most clinically-unidentified DLB subjects, who also have concurrent neuropathology-criteria AD (AD/DLB), as well as those with neuropathological AD and amygdala-predominant LBD insufficient for a DLB diagnosis, may best be classified as having a “secondary synucleinopathy. Importantly, the DLB Consortium recognized the importance of comorbid AD pathology by defining “Low”, “Intermediate” and “High” subdivisions of DLB based on the relative brain stages of both Lewy body and AD pathology. If the one-year rule for separating PD from DLB, and for then dividing DLB into subtypes based on the presence and severity of comorbid AD pathology, is effective, then the divided groups should statistically differ in important ways. In this study we used the comprehensive clinicopathological database of the Arizona Study of Aging and Neurodegenerative Disorders (AZSAND) to empirically test this hypothesis. Furthermore, we used multivariable statistical models to test the hypothesis that comorbid AD neuropathology is a major predictor of the presence and severity of postmortem Lewy synucleinopathy. The results confirm the clinicopathological heterogeneity of Lewy body disorders as well as the profound influence of comorbid AD pathology.

## INTRODUCTION

In recent years, proposals have been advanced to redefine or reclassify Lewy body disorders by merging the long-established entities of Parkinson’s disease (PD), Parkinson’s disease dementia (PDD) and dementia with Lewy bodies (DLB). Some (1-3) have based their argument on clinical and genetic findings that “… challenge the central role of the classical pathologic criteria” and other groups (4,5) argue from the opposite side, on the basis that these conditions are alpha-synucleinopathies and that other differences are secondary, while another new system continues to recognize the heterogeneity of Lewy body diseases (6). All propose new, biomarker-based clinical classifications.

We here advance a critique of the proposed unity of sporadic, late-onset PD and DLB and point out in particular the probable shortcomings of these proposed recent re-classification approaches. The International DLB Consortium classification system (7-11), evolved over almost three decades of consensus collaborations between neurologists, neuropsychologists and neuropathologists, has generated a wealth of clinicopathological studies (12-35) that have very convincingly established that PD and DLB differ in many ways that would greatly complicate unitary clinical trials by introducing tremendous subject heterogeneity. While the “one year rule” for separating PD and DLB has been criticized as arbitrary, it has been a pragmatic and effective tool for splitting the continuum between the two entities. Neuropathologists have long recognized at least two distinct underlying patterns of synuclein pathology spread and these have been confirmed by data-driven, autopsy-based statistical clustering analyses (36-38).

In addition to the decades of literature supporting the non-homogeneity of PD and DLB, it has become increasingly apparent that Lewy body disorders may fundamentally differ in their etiology. Most PD subjects, as well as most clinically-presenting DLB subjects, might best be classified as having a “primary synucleinopathy” while most clinically-unidentified DLB subjects, who also have concurrent neuropathology-criteria AD (AD/DLB), as well as those with neuropathological AD and amygdala-predominant LBD insufficient for a DLB diagnosis, may best be classified as having a “secondary synucleinopathy” (39-45). This is analogous to the accepted classification of tauopathies into primary and secondary categories (46-49). Both tau and synuclein pathology clearly occur as responses to several different inherited cerebral amyloidoses, including genetic early-onset forms depositing not only Aβ (point mutations in *PSEN* and *APP*, trisomy 21) but also prion (Gerstmann-Straussler-Scheinker disease) and gelsolin variant proteins (50-63). Also, synuclein pathology accompanies 60-70% of sporadic AD, even in those subjects with very early onset (64) and, in the Arizona Study of Aging and Neurodegenerative Disorders (AZSAND), roughly 35% and 50% of clinicopathologically-diagnosed PD and Parkinson’s disease dementia (PDD) cases have neuropathology-criteria AD, respectively. This evidence suggests that the majority of human synuclein pathology may be secondary to Aβ cerebral amyloidosis.

The DLB Consortium recognized the importance of comorbid AD pathology by defining “Low”, “Intermediate” and “High” subdivisions of DLB based on the relative brain stages of both Lewy body and AD pathology. If the one-year rule for separating PD from DLB, and for then dividing DLB into subtypes based on the presence and severity of comorbid AD pathology, is effective, then the divided groups should statistically differ in important ways. In this study we used the comprehensive AZSAND clinicopathological database to empirically test this hypothesis. Furthermore, we used multivariable statistical models to test the hypothesis that comorbid AD neuropathology is a major predictor of the presence and severity of postmortem Lewy synucleinopathy.

## METHODS

### Subject selection

Subjects were selected by database searches of the AZSAND/Brain and Body Donation Program (www.brainandbodydonationprogram.org) (65). Search criteria specified that subjects died and had a complete neuropathological examination with clinicopathological diagnoses of PD, DLB, or AD, or were non-demented, non-parkinsonian controls with or without Lewy pathology. Selected DLB subjects had dementia and met intermediate or high likelihood neuropathological criteria for DLB (9,11). Selected AD subjects had dementia and met intermediate or high National Institute on Aging-Reagan Institute (NIA-RI) and/or NIA-Alzheimer’s Association AD neuropathological criteria (66,67). AD subjects not also diagnosed with PD and not meeting intermediate or high DLB likelihood were classified as Alzheimer’s disease with Lewy bodies (ADLB)(15,65). Selected PD subjects met AZSAND PD criteria, including clinical parkinsonism with substantia nigra neuron loss and Lewy pathology. Intermediate and high NIA-RI/NIA-AA AD criteria (66,67) stipulate Braak neurofibrillary stages III or IV versus V and VI, respectively, with moderate or frequent neuritic plaques. DLB intermediate and high criteria are based on comparison of Lewy body pathology stage with AD pathology stage; when AD NIA level is high, only the neocortical Lewy body stage qualifies for DLB, while when it is intermediate, either a limbic or neocortical Lewy body stage qualifies for DLB.

### Subject characterization

Most subjects had serial standardized research-dedicated clinical evaluations, done by teams of nurses, medical assistants, behavioral neurologists, movement disorders neurologists, neuropsychologists and psychometrists using standardized assessment batteries [64], including the Mini Mental State Examination (MMSE), National Alzheimer’s Coordinating Center (NACC) Uniform Data Set (UDS) and the Unified Parkinson’s Disease Rating Scale (UPDRS). Subjects had olfactory testing with the University of Pennsylvania Smell Identification Test (67)(UPSIT) every third year on average.

All subjects received identical neuropathological examinations by a single observer (TGB), including summary regional brain density measures for total amyloid plaques and neurofibrillary tangles (summary score of 5 regional semi-quantitative 0-3 density scores for a maximum possible total of 15 in frontal, temporal and parietal lobes plus hippocampal CA1 and entorhinal/transentorhinal area), Lewy body pathology summary regional brain density scores (summary score of semi-quantitative 0-4 density scores in 10 brain regions for a maximum possible total of 40), and staging using the Unified Staging System for Lewy Body Disorders [15], as well as assignment of CERAD neuritic plaque density, Braak neurofibrillary stage, and AD neuropathological change levels of Low, Intermediate or High, as described previously [65].

### Statistical analysis

Demographic and post-mortem characteristics were analyzed using one-way analysis of variance (ANOVA), Chi-square tests, Fisher’s Exact tests and paired significance testing as appropriate. The objectives were twofold, first to determine whether the subgroups were significantly different in terms of age of disease onset, disease duration, fraction that were male, percentage meeting AD neuropathological criteria (NIA-R/NIA-AA intermediate or high), fraction with the apolipoprotein E-ℰ4 allele, fraction with severe substantia nigra pigmented neuron loss, fraction with clinical parkinsonism, and severity scores for UPSIT, UPDRS motor score, MMSE score and summary brain synuclein pathology load, and second, to determine, using logistic regression models adjusted for age, gender and possession of an apolipoprotein E-ℰ4 allele, the independent influence of AD neuropathology on the presence and severity of brain synuclein pathology.

## Results

Clinical, demographic and neuropathological characteristics of the compared groups are shown in Table 1 and Figure 1. Analysis of variance or chi-square tests found significant group differences on all comparisons. Despite multiple specific paired differences between subgroups (see Supplementary Data File 1 for raw data and complete statistical results), on most comparisons the diagnostic groups fell into two distinct sets that were collectively different, as shown with red and blue coloration of the bar graphs, respectively. The red subgroups appear to be most reflective of synuclein pathology while the blue subgroups are more influenced by AD pathology. The PD and PDAD groups are distinctly different from all other groups in age of onset (youngest, Figure 1a), disease duration (longest, Figure 1b), percentage with parkinsonism (most, Figure 1g), percentage with severe SN neuronal loss (most, Figure 1h), UPDRS motor score (highest, Figure 1i) and MMSE score (highest, Figure 1f). For other characteristics, one or more of the DLB groups are distinctly similar to the PD groups, including for percentage male (most, Figure 1c), UPSIT score (lowest, Figure 1l), brain synuclein pathology load (most, 1j) and percentage with peripheral nervous system (PNS) synuclein pathology (most, 1k).

**Table 1.** Diagnostic group sizes and mean ages. PD = PD (non-demented) + PDD (with dementia); DLB-H = DLB Consortium High Likelihood of DLB; DLB-I = DLB Consortium Intermediate Likelihood of DLB; ADLB = AD with DLB Consortium Low Likelihood Lewy body pathology; ADNLB = AD with no Lewy body pathology. Means and standard deviations are shown. See also Supplementary Files 1 and 2.

|  | PD<sup>1</sup><br>(n = 179) | PDAD<br>(n = 90) | DLB-H<br>(n = 65 ) | DLB-I<br>(n = 91 ) | ADLB<br>(n = 306 ) | ADNLB<br>(n = 421 ) | ANOVA p-value |
| --- | --- | --- | --- | --- | --- | --- | --- |
| Age (yrs) | 79.55<br>(7.14) | 81.56<br>(6.13) | 81.78<br>(6.66) | 82.97<br>(9.16) | 82.62<br>(9.57) | 84.02<br>(9.57) | < 0.0001 |

1. PD significantly different from DLB-I (Tukey $p > 0.001$ ); PD significantly different from ADLB and ADNLB (Tukey $p > 0.0001$ )

**Figure 1.**
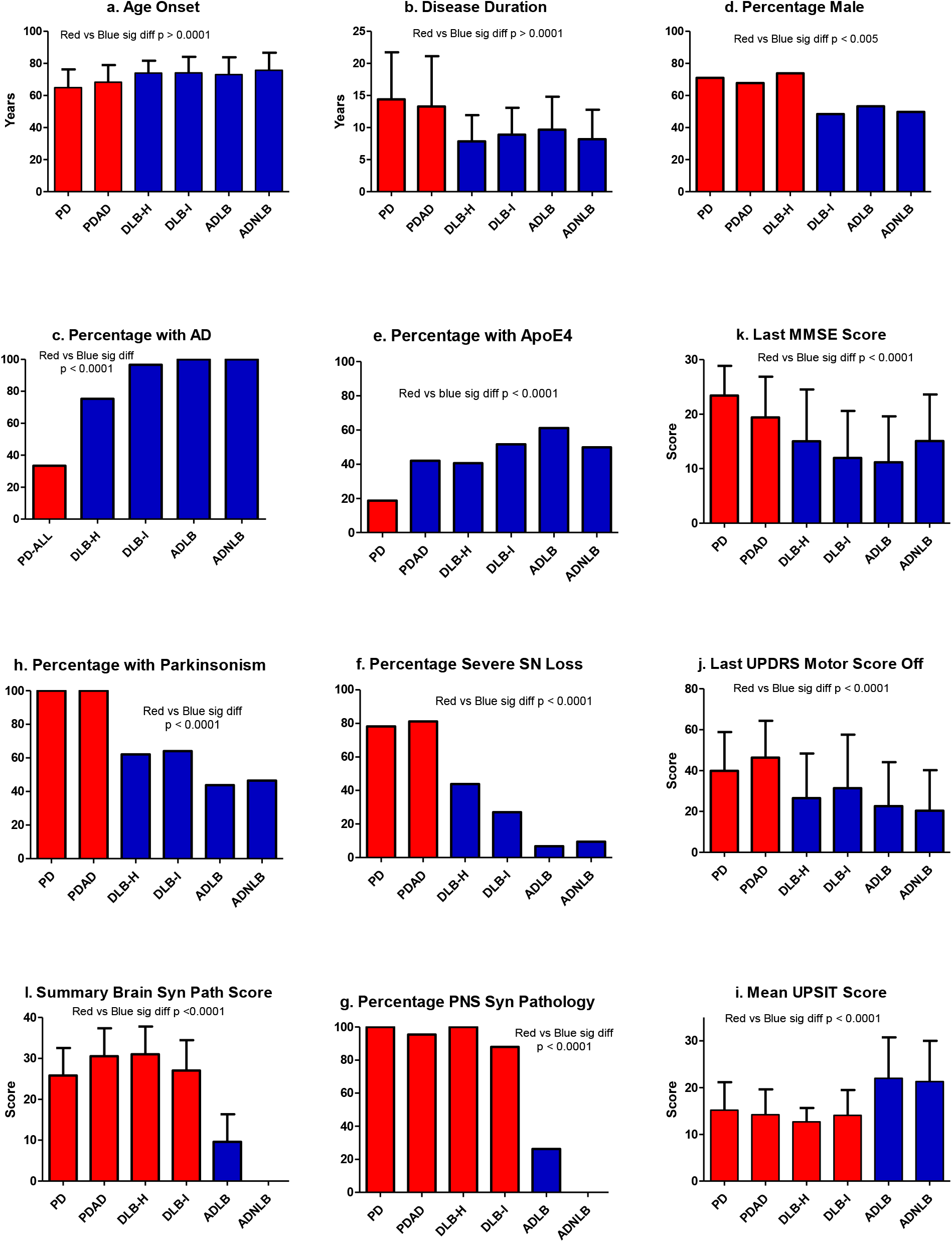
Comparison of clinicopathological subgroups defined by PD, DLB and AD criteria of the International Dementia with Lewy Bodies Consortium and the National Institute on Aging-Reagan/Alzheimer’s Association. See Supplementary File 1 for complete data and statistical subgroup comparisons. The subgroups colored red appear to be most influenced by synuclein pathology while those colored blue are most influenced by AD pathology. PNS = peripheral nervous system.

Logistic regression analysis of AZSAND data (Supplementary File 2) shows that the presence of threshold brain distributions of amyloid plaques are the strongest predictor, with a 1.49 Odds Ratio (OR) and p-value < 0.0001, for the presence of any brain synuclein pathology (Table 2), exceeding the association strengths of age, male sex and the apolipoprotein E-ℰ4 allele. For subjects clinicopathologically diagnosed with PD (Table 3), threshold levels of both plaques and tangles are again the strongest predictors, with 2.7 and 2.4 ORs, respectively, for the presence of the neocortical stage of Lewy synuclein pathology, the most severe form of Lewy body disease, again exceeding the previously-known association strengths of age, male sex and the apolipoprotein E-ℰ4 allele. The plaque and tangle load thresholds for both regressions were chosen to approximate the thresholds at which PET-tau and PET-amyloid become positive.

**Table 2.** Univariable and multivariable logistic regression models assessing predictors for the presence of any brain Lewy synuclein pathology (see Supplementary File 2 for data).

| Variable | Effect | Univariable |  | Multivariable |  |
| --- | --- | --- | --- | --- | --- |
|  |  | OR (95% CI) | P value | OR (95% CI) | P value |
| **Age** | One year change | 0.99 (0.98,1) | 0.016 | 0.99 (0.98,1) | 0.031 |
| **Sex** | Male vs Female | 1.39 (1.15,1.69) | < 0.001 | 1.41 (1.16,1.72) | < 0.001 |
| **ApoE4 any** | Yes vs No | 1.6 (1.31,1.95) | < 0.001 | 1.31 (1.06,1.63) | 0.014 |
| **Tangle score > 5** | Yes vs No | 1.28 (1.05,1.56) | 0.016 | 1.11 (0.89,1.4) | 0.349 |
| **Plaque score > 5** | Yes vs No | 1.64 (1.34,2) | < 0.001 | 1.49 (1.18,1.9) | < 0.001 |

**Table 3.** Univariable and multivariable logistic regression model assessing predictors for the presence, in subjects clinicopathologically diagnosed with Parkinson’s disease, of the Neocortical Stage of brain Lewy synuclein pathology (see Supplementary File 2 for data).

| Variable | Effect | Univariable |  | Multivariable |  |
| --- | --- | --- | --- | --- | --- |
|  |  | OR (95% CI) | P value | OR (95% CI) | P value |
| **Age** | One year change | 1.02 (0.98,1.06) | 0.347 | 1 (0.96,1.05) | 0.924 |
| **Sex** | Male vs Female | 1.08 (0.61,1.88) | 0.798 | 1.34 (0.72,2.49) | 0.351 |
| **ApoE4 any** | Yes vs No | 2.68 (1.43,5.19) | 0.003 | 1.96 (0.96,4.01) | 0.066 |
| **Tangle score > 5** | Yes vs No | 2.76 (1.59,4.89) | < 0.001 | 2.42 (1.32,4.43) | 0.004 |
| **Plaque score > 5** | Yes vs No | 3.73 (2.16,6.55) | < 0.001 | 2.77 (1.5,5.11) | 0.001 |

## DISCUSSION

The apparent dichotomy of results for the diagnostic subgroups, albeit with variable dyadic constituents, supports to some extent the division of Lewy body diseases into primary and secondary synucleinopathies, with primary synucleinopathies composed of PD and PDAD, while DLB-H, DLB-I, and ADLB might be considered secondary synucleinopathies due to their greater similarities, in several categories, to AD without Lewy pathology (ADNLB), including older age of onset, shorter disease duration, greater fraction with an apolipoprotein E-ℰ4 allele, lower final MMSE and UPRS motor scores, and smaller fractions with parkinsonism and severe SN neuronal loss. However, for some other categories the DLB-H and/or DLB-I subgroups are more similar to PD and PDAD, including for greater fraction of males affected, lower UPSIT olfactory scores, and greater CNS and PNS synuclein pathology scores, suggesting that these subgroups may be more or less equally influenced by synuclein and AD pathology. It is possible that synuclein and AD molecular pathologies may have separate and independent origins with a subsequent acceleration of the prion-like spread of synucleinopathy in subjects with comorbid AD. These very distinct “biological” subgroup differences indicate that PD and most cases of DLB will need completely different approaches to clinical diagnosis, prevention and therapy and therefore will also need continued efforts to more fully explore differences, as well as similarities, in their etiology and pathogenesis.

Our conclusion thus runs counter to what has been envisioned for PD and DLB classifications in recent publications (1-5), where statements have included, “Disease staging is a classification system that produces clusters of patients who require similar treatments and have similar expected outcomes. Staging can serve as the basis for clustering clinically homogeneous patients”. In fact, we believe that staging as proposed, by lumping PD and DLB together, rather than decreasing heterogeneity of the grouped subjects, would increase it. This is in opposition to the general direction across medical fields towards “personalized” or “precision” medicine and would hinder potential trial-enhancing stratification of patients in clinical trials.

Note that the clinicopathological heterogeneity here demonstrated would likely be further increased when considering additional neuropathological comorbidities including microscopic changes of progressive supranuclear palsy, amyloid angiopathy, TPD-43 proteinopathy, cerebrovascular ischemic lesions, ARTAG, argyrophilic grains and others.

We therefore support new synuclein biomarker-based clinical classifications of Lewy body disorders but propose that any new classificatory systems should be set within the established clinicopathological context principally developed by the International DLB Consortium. Aided by biomarker identification of both AD and synuclein molecular pathology, cross-referencing the newly-proposed classificatory categories with the extensively documented DLB and NIA-AA AD criteria will allow drawing on the decades-long published clinicopathological literature for both Lewy body disorders and Alzheimer’s disease. Concurrent biomarker assays for pathology associated with LBDs and AD will now make such cross-referencing possible and should enable enhanced preclinical detection of LBD as well as improved prediction of rates of cognitive and motor decline. With the new wave of greatly increased biomarker diagnostic accuracy, recognition of the often clinically unsuspected presence of AD within subjects clinically diagnosed with PD and DLB, as well as the clinically unsuspected presence of Lewy body disease in subjects clinically diagnosed with AD, will allow for eventual clinical trials directed at molecular therapy for two or even all three molecular targets simultaneously (e.g. Aβ, tau and synuclein), an eventuality that seems increasingly necessary.

## Data Availability

All data produced in the present work are contained in the manuscript and Supplemental Files.

## ACKNOWLEDGEMENTS

The Brain and Body Donation Program has been supported by the National Institute of Neurological Disorders and Stroke (U24 NS072026 National Brain and Tissue Resource for Parkinson’s Disease and Related Disorders), the National Institute on Aging (P30 AG019610 and P30AG072980, Arizona Alzheimer’s Disease Core Center), the Arizona Department of Health Services (contract 211002, Arizona Alzheimer’s Research Center), the Arizona Biomedical Research Commission (contracts 4001, 0011, 05-901 and 1001 to the Arizona Parkinson’s Disease Consortium) and the Michael J. Fox Foundation for Parkinson’s Research.

